# Vaccination in a two-group epidemic model

**DOI:** 10.1101/2021.01.10.21249557

**Authors:** Sebastian Aniţa, Malay Banerjee, Samiran Ghosh, Vitaly Volpert

## Abstract

Epidemic progression depends on the structure of the population. We study a two-group epidemic model with the difference between the groups determined by the rate of disease transmission. The basic reproduction number, the maximal and the total number of infected individuals are characterized by the proportion between the groups. We consider different vaccination strategies and determine the outcome of the vaccination campaign depending on the distribution of vaccinated individuals between the groups.

## 1 Introduction

Epidemic progression in a heterogeneous population depends on the proportion of different groups characterized by the rate of disease transmission [3, 4, 5, 6]. In this work we study the efficacy of vaccination campaign for different vaccination strategies. We consider the epidemic progression in a two-group population consisting of susceptible (*S*_1_, *S*_2_) and infected (*I*_1_, *I*_2_) compartments and described by the following model:

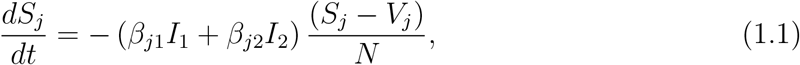

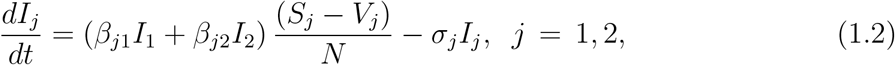

where *β*_*ij*_ are the rates of disease transmissions, *σ*_*j*_ (*j* = 1, 2) are the clearance rates of infected individuals, and *N* is the total population. Here *V*_1_ and *V*_2_ denote the number of vaccinated in the first and second group, respectively. Let us note that the number of individuals in each class, which can be infected, is *S*_*j*_ − *V*_*j*_, *j* = 1, 2. We suppose that vaccination is fully efficient in the sense that vaccinated individuals cannot become infected.

For this model we are intended to calculate the basic reproduction number, size of the epidemic and study the effect of vaccination to arrest the disease progression.

## 2 Epidemic indicators

In this section we determine the basic reproduction number in the heterogeneous population, the total and the maximal number of infected individuals depending on the number of vaccinated individuals.

### 2.1 Basic reproduction number

At the beginning of epidemic, let us assume that *S*_10_ and *S*_20_ denote the number of susceptible individuals in the two groups such that *S*_10_ + *S*_20_ = *N*, and we define 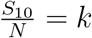 and 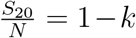, where 0 ≤ *k* ≤ 1. The Jacobian matrix of the system (1.1) - (1.2) evaluated at the disease free equilibrium point is

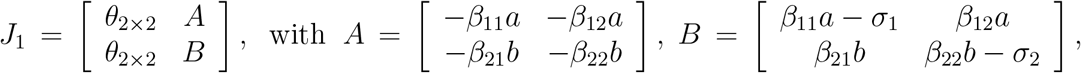

with 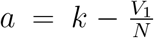 and 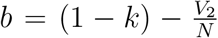. Clearly, two eigenvalues equal to zero, while the largest eigenvalue is given by

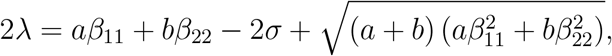

where we have assumed *σ*_1_ = *σ*_2_ = *σ, β*_12_ = *β*_21_ = (*β*_11_ +*β*_22_)*/*2 for simplicity of presentation. Equating the largest eigenvalue to zero, we find the basic reproduction number

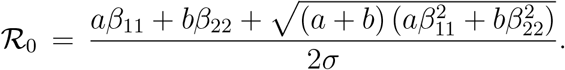

### 2.2 Region of epidemic growth

Epidemic growth occurs for ℛ_0_ *>* 1. Depending on the number of vaccinated individuals in the two classes, we can determine the regions in the (*V*_1_, *V*_2_)-parameter plane, where the epidemic progresses. Since the number of vaccinated in each group cannot be greater than the initial number of susceptible, then *V*_1_ ≤ *kN* and *V*_2_ ≤ (1 − *k*)*N*. The boundary between the regions of epidemic growth and the region epidemic extinction can be obtained from the relation *ℛ* _0_ = 1:

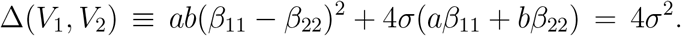

The epidemic extinction occurs in *E*_1_ and the epidemic progresses in *E*_2_ where:

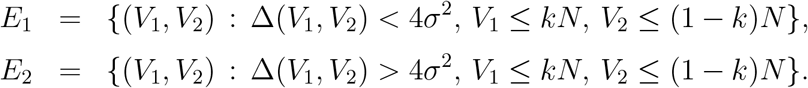

Fig. 1 shows the regions with epidemic growth and extinction for different values of *k*.

**Figure 1:**
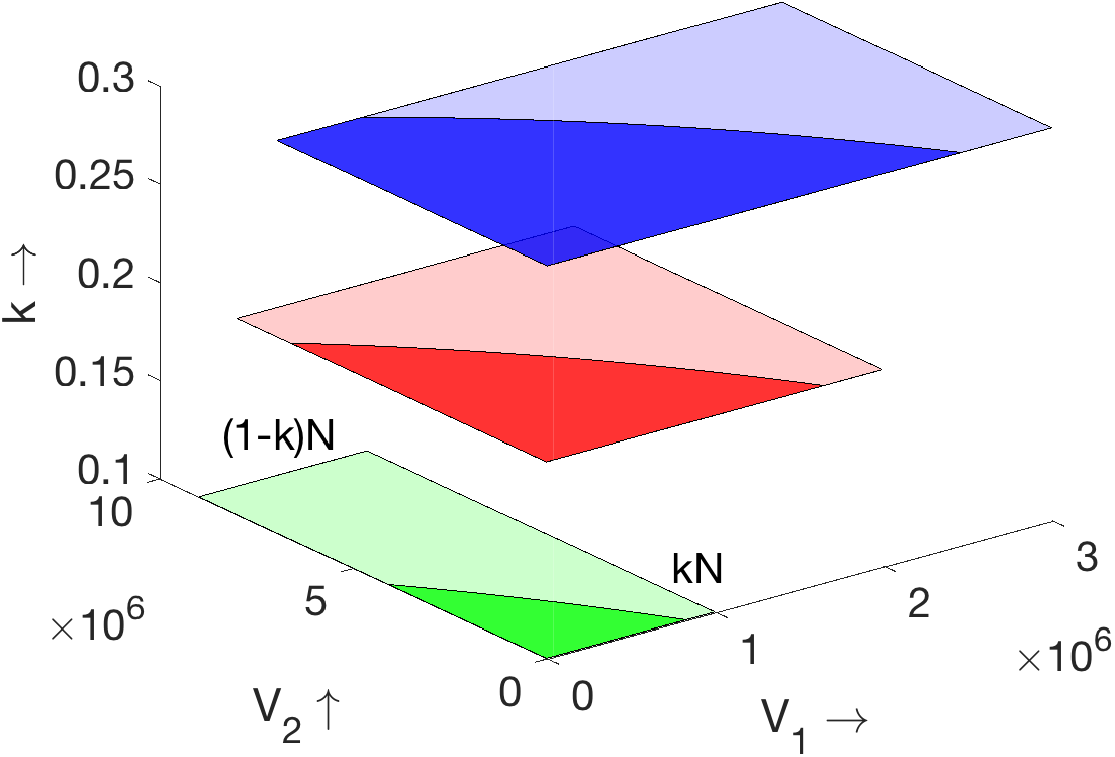
Regions of epidemic growth and extinction are marked with deep and light colours, respectively. Parameter values are *β*_11_ = 4, *β*_22_ = 1, *β*_12_ = *β*_21_ = 2.5, *σ*_1_ = *σ*_2_ = 1 and *k* = 0.1 (green), *k* = 0.2 (red) and *k* = 0.3 (blue).

### 2.3 Final size of epidemic

Taking a sum of equations (1.1), (1.2), and then integrating between *t* = 0 and *t* = ∞, we obtain the equalities:

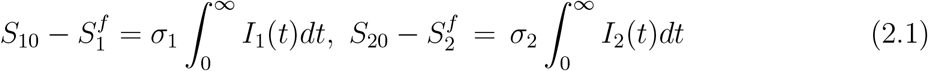

under the assumption that *I*_*j*_(0) = *I*_*j*_(∞) = 0, *j* = 1, 2 and 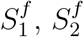 are the final sizes of two susceptible groups. Next, we divide equation (1.1) by *S*_*j*_, and integrate from 0 to ∞:

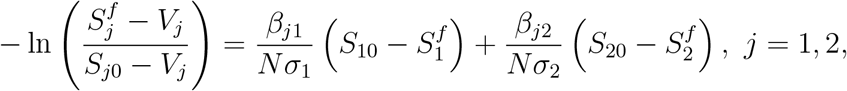

using the equalities in (2.1). Introducing four new quantities 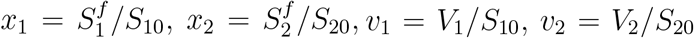, and assuming that *σ*_1_ = *σ*_2_ =*σ, β*_12_ = *β*_21_ = (*β*_11_ + *β*_22_)*/*2, we obtain the following equations

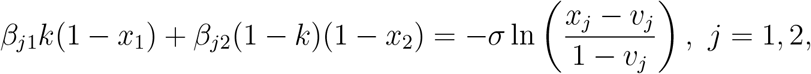

with respect to *x*_1_ and *x*_2_. The positive solution of this system, satisfying the restriction 0 *< x*_1_, *x*_2_ *<* 1, determines the final size of susceptible populations (Fig. 2, left). We can now determine the number of infected individuals in each group at the end of epidemic and the total number of infected in both groups, 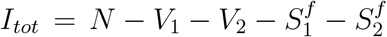.

**Figure 2:**
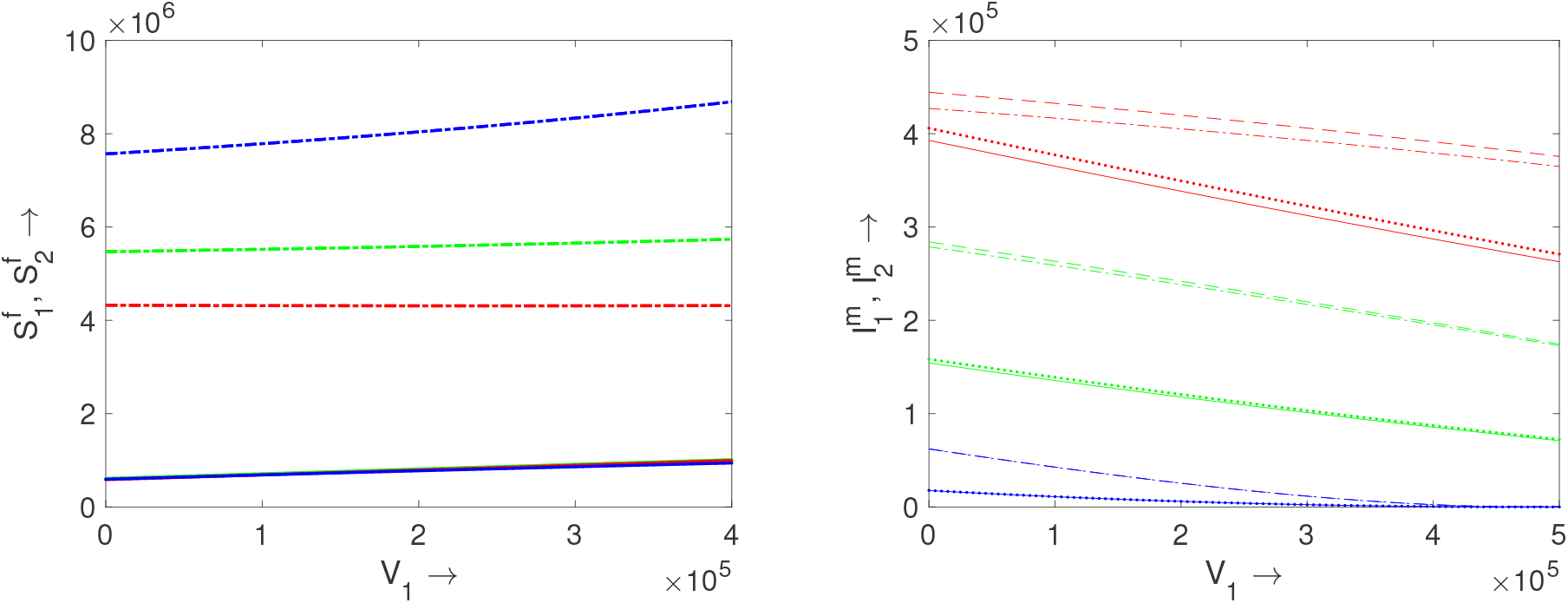
Left panel: analytical and numerical values of the final sizes of epidemic for two groups for *k* = 0.1 (blue), *k* = 0.2 (green), *k* = 0.3 (red). Right panel: analytical and numerical values of maximum number of infected individuals for two groups for *k* = 0.1 (blue), *k* = 0.2 (green), *k* = 0.3 (red). Other parameter values are as follows: *β*_11_ = 4,*β*_22_ = 1, *β*_12_ = *β*_21_ = 2.5, *σ* = 1, *V*_2_ = 2.4 · 10^6^ − *V*_1_.

### 2.4 Maximum number of infected

In order to find the maximal number of infected individuals in the heterogeneous population, we consider an approximation 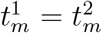, where 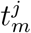 is time at which the functions *I*_*j*_(*t*) reach their maxima (*j* = 1, 2). Integrating equations 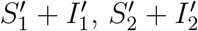 from 0 to *t*_*m*_, we obtain:

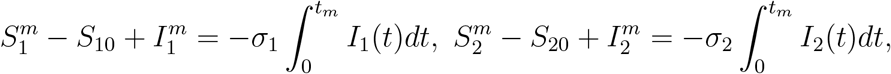

where 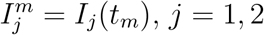. Next, we divide equation (1.1) by *S*_*j*_, and integrate over [0, *t*_*m*_] to find:

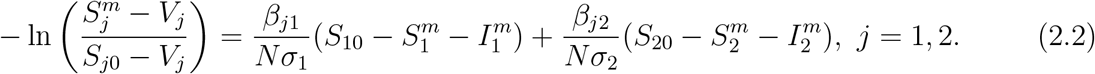

Assuming that 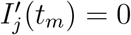, we get from (1.2):

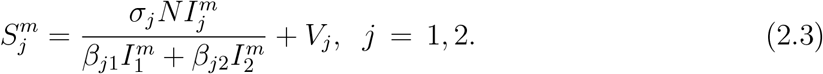

As before, we assume that *σ*_1_ = *σ*_2_ = *σ* and set 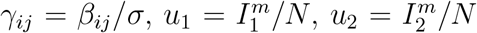, and *v*_1_, *v*_2_ as defined in the previous subsection. Using (2.3), we can rewrite equation (2.2) as follows:

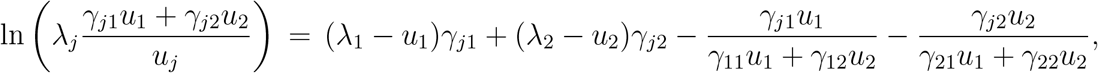

where *λ*_*j*_ = ((*j* − 1) + (3 − 2*j*)*k*)(1 − *v*_*j*_), *j* = 1, 2. Solving this system of equations, we find *u*_*j*_ and, consequently, 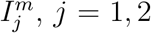 (Fig. 2, right). We then use formulas (2.3) to determine 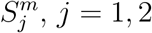.

## 3 Optimization of vaccination strategies

Since there is a cost related to vaccination, an optimal control problem (optimal vaccination) is proposed:

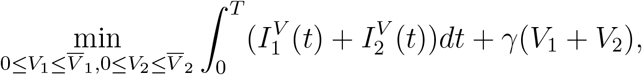

meaning that we are interested to minimize total damage of the epidemics, which includes the cost related to the total number of infected individuals and the cost of the vaccine. Here, 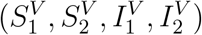 is the solution to (1.1)-(1.2) which satisfies the initial conditions 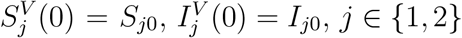, where 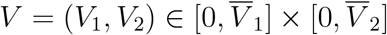 is the control/vaccination strategy. Since, due to certain constraints, not all individuals may be vaccinated we have imposed the constraints on 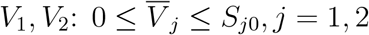 and *γ* is a positive constant. Let

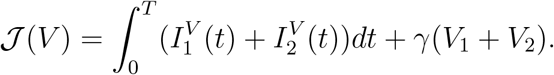

Consider the following adjoint problem

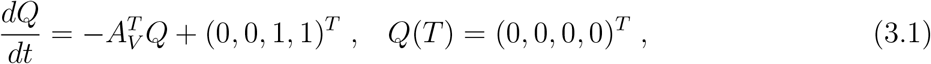

where 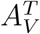 is the transposed of the matrix

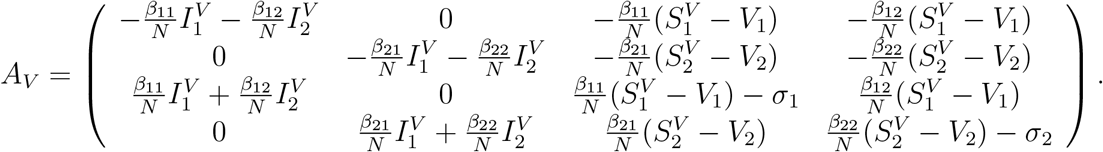

Let *Q*^*V*^ be a solution of problem (3.1), 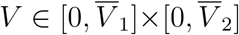 and 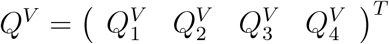. Following the method presented in [1, 2], we can prove that for any *θ* ∈ ℝ^2^ such that 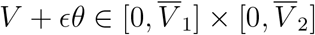 for any sufficiently small *ϵ >* 0.

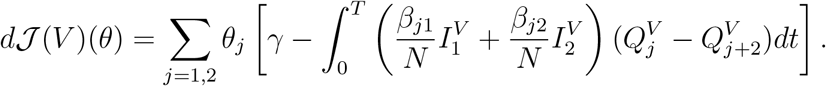

The evaluation of directional derivative of 𝒥 allows us to derive a conceptual iterative algorithm (gradient type) which improves at each step the control *V* = (*V*_1_, *V*_2_). On the other hand, we can prove as in [1] that there exists at least one optimal control 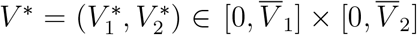 (optimal vaccination strategy), *i*.*e*., 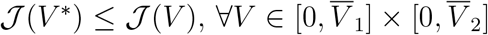.

Using the form of the directional derivative of 𝒥 we deduce that

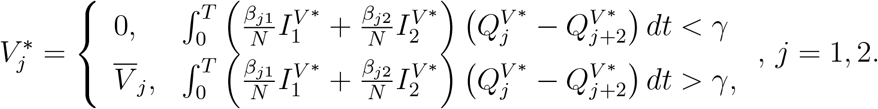

This allows us to improve the above mentioned gradient-type algorithm. More detailed analysis and applications of this approach will be presented elsewhere.

Different vaccination strategies are illustrated in Fig. 3. Each point on the (*V*_1_, *V*_2_)-plane shows one vaccination with the corresponding numbers of vaccinated individuals in each class. As such, for the first vaccination we have *V*_1_ + *V*_2_ = 10^6^ for all vaccination strategies but the proportion between the two groups are different. Similarly, the total number of vaccinated individuals equals 2 · 10^6^ after the second vaccination, and so on. Thus, we consider the same total number of vaccinated individuals at each vaccination stage with different proportions between the classes in different vaccination strategies.

**Figure 3:**
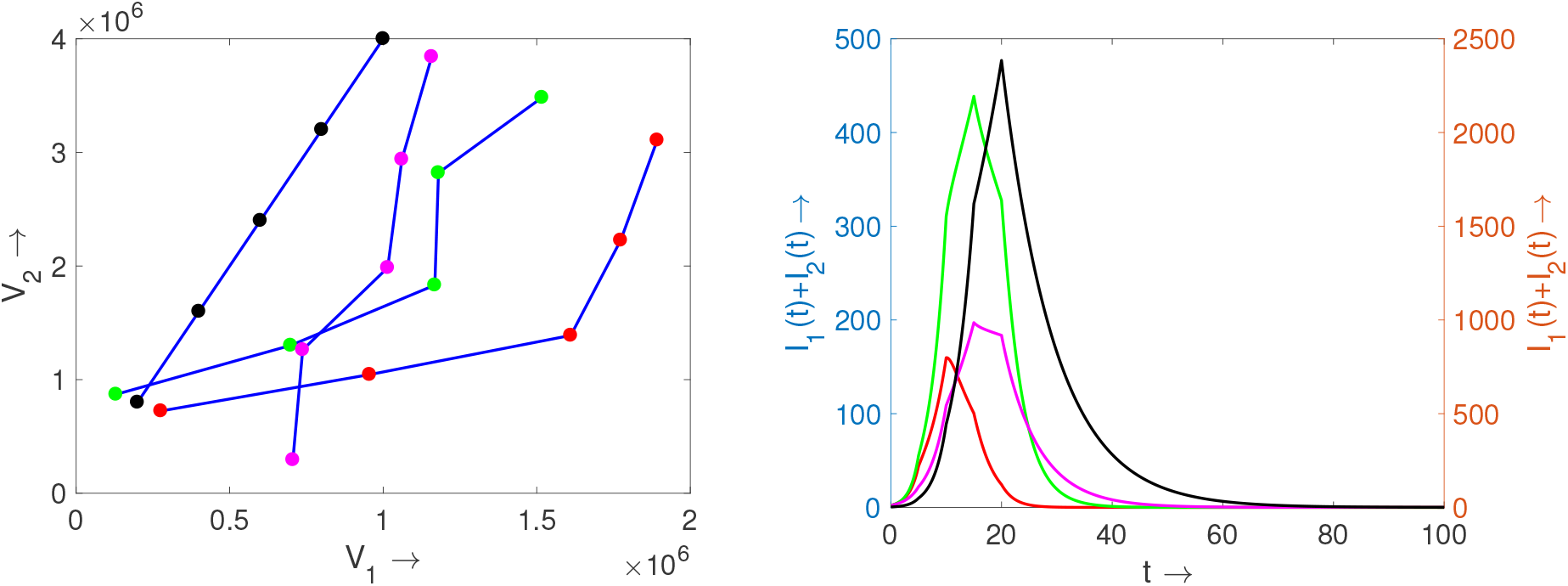
Left panel: various vaccination strategies shown as lines in the (*V*_1_, *V*_2_)-plane. Right panel: plots of *I*_1_(*t*)+ *I*_2_(*t*) for four vaccination strategies shown in the left figure. The colours of the curves correspond to the colours of respective dots in the left panel. Scaling of the black curve is shown at the right vertical axis and of other curves the left vertical axis. Parameter values are the same as in Fig. 2.

The curve with black dots in the left panel of Fig. 3 corresponds to a random choice of vaccinated individuals. Since for *k* = 0.2 we have *S*_2_*/S*_1_ = (1 − *k*)*/k* = 4, then setting *V*_2_ = 4*V*_1_ we obtain the same proportion of vaccinated individuals in each class as the proportion between the classes of susceptible individuals. We neglect here the change of *S*_2_*/S*_1_ during the epidemic progression. Though this vaccination strategy is quite natural, it is less efficient compared to the other strategies where more individuals from *S*_1_ are vaccinated.

A total number 5 · 10^6^ of individuals are vaccinated in five times with 10^6^ vaccines each one. The first vaccination is effectuated at the beginning of the (*t* = 0) and the remaining vaccinations after every five time units. For various vaccination strategies shown in Fig. 3 (left), the corresponding functions *I*(*t*) = *I*_1_(*t*) + *I*_2_(*t*) are shown in Fig. 3 (right). We can characterize the vaccination strategies by the functions *V*_2_(*t*) = *f*_*i*_(*V*_1_(*t*)), *i* = 1, 2, 3, 4. We conclude that for any two vaccination strategies *f*_1_ and *f*_2_ such that *f*_1_(*V*_1_) *< f*_2_(*V*_1_), the corresponding functions (sum of infected individuals in both classes) satisfy the inequality *I*^1^(*t*) *< I*^2^(*t*) for all *t* > 0. A similar relation holds for the total number of infected individuals at the end of epidemic.

Thus, vaccination of the first class of susceptible individuals is more efficient from the point of view of minimizing the number of infected individuals. This conclusion can be expected because the rate of disease transmission by this group is faster. However, this conclusion may not hold true if we minimize the number of deaths taking into account different mortality rates in the two groups. Let us consider an example with two groups: *S*_1_ corresponds to people less than 60 years old, *S*_2_ to people more than 60. Assume that *k* = 0.8, that is, *S*_10_ = 0.8*N* and *S*_20_ = 0.2*N*. Consider the vaccination strategies where *V*_1_*/V*_2_ =const and let us vary this ratio. The total numbers of infected individuals in each group at the end of epidemic are presented in the following table.

| $V_1/V_2$ | $V_1/V_2 = 8$ | $V_1/V_2 = 4$ | $V_1/V_2 = 1.5$ |
| --- | --- | --- | --- |
| $I_1^{tot}$ | 5 476 432 | 5 707 105 | 6 209 094 |
| $I_2^{tot}$ | 1 364 707 | 1 227 671 | 908 982 |

The ratio *V*_1_*/V*_2_ = 4 corresponds to a random choice of vaccinees. As before, increase of the proportion of the first group decreases the total number of infected 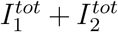. However, the total number of deaths depends on the mortality rate in each group. In the application to the Covid-19, we assume that the mortality rate of infected individuals in the second group is of the order of magnitude 10 times larger than in the first group. In this case, the total number of deaths decreases for a larger proportion of vaccinees in the second group.

## Data Availability

No data used.

## Acknowledgements

The last author has been supported by the RUDN University Strategic Academic Leadership Program.

